# Deep brain stimulation-responsive subthalamo-cortical coupling in obsessive-compulsive disorder

**DOI:** 10.1101/2025.06.12.25329123

**Authors:** Lucie Winkler, Lucy M. Werner, Markus Butz, Christian J. Hartmann, Alfons Schnitzler, Jan Hirschmann

## Abstract

Deep brain stimulation (DBS)-responsive oscillations have been implicated in motor symptoms of Parkinson’s disease (PD). Their role in non-movement disorders, such as obsessive-compulsive disorder (OCD), is less clear. Here, we aimed to characterize the effect of DBS on subthalamic and cortical oscillations in OCD.

Local field potential recordings from the subthalamic nucleus (STN) were combined with magnetoencephalography in one OCD patient at rest (DBS OFF and ON) and in a Go/NoGo task (DBS OFF). A PD patient completed the same task for comparison.

In the OCD patient, we observed right-lateralized beta peaks in STN power and STN-cortex coherence. These were diminished by DBS. Task-related modulations of STN power occurred in the theta band for the OCD patient, and in the beta band for the PD patient.

We conclude that resting-state, DBS-responsive beta oscillations are not necessarily a sign of Parkinsonism. Task-related spectral modulations might be more disease-specific than resting-state oscillations.

## Introduction

Beta oscillations within the subthalamic nucleus (STN) are central to understanding the pathophysiology of Parkinson’s disease (PD) and the therapeutic mechanisms of deep brain stimulation (DBS). In PD patients, beta activity is pathologically enhanced in the STN and other structures of the cortico-basal ganglia loop ^1–3^, and is widely recognized to contribute to PD motor symptoms such as bradykinesia and rigidity ^4,5^. DBS of the STN alleviates motor symptoms, presumably by reducing excessive beta activity in the STN ^6–8^ and sensorimotor cortex ^6,9^. These findings suggest a causal role of subthalamic beta oscillations in motor slowing. This notion, however, is mostly based on observations in PD patients.

Here, we examined a patient with obsessive-compulsive disorder (OCD), a psychiatric condition marked by persistent thoughts and urges (obsessions), and repetitive actions or mental operations (compulsions) ^10^. OCD is characterized by pathologically enhanced overconnectivity in a network spanning limbic cortical regions, the striatum, and the STN ^11–15^. Altered theta activity in the STN ^16,17^ and cortex ^18–20^ has been identified as a potential biomarker of OCD pathology.

Importantly, OCD is a condition for which, despite the absence of motor slowing, DBS of the STN is being applied as a therapeutic intervention. The therapeutic benefit possibly acts through a reduction of theta oscillations in the fronto-basal ganglia pathway ^21,22^. The role of beta oscillations in OCD has not been studied extensively, but some studies suggest that beta activity is altered in cortex ^23^ and the STN ^16^ and that DBS is associated with both increases ^22^ and decreases ^24^ of beta activity in the stria terminalis/anterior limb of the internal capsule and frontal cortex. However, while it has been demonstrated that beta oscillations are present in the dorsal ^17^ and anteromedial STN ^11^ in OCD, it remains unknown whether and how DBS influences these oscillations. Therefore, we aimed to examine the effect of DBS in OCD.

## Materials and methods

### Patients

A female patient in her fifties suffering from severe OCD (first manifestation in the third decade), marked by excessive washing of the hands, participated in the present study. She was implanted with DBS electrodes (3389) 12 years before measurement, and received a new stimulator one day before participating in the present study. DBS reduced her Yale-Brown Compulsive Obsessive Scale score substantially, from 39/40 pre-operatively to 7 at the time of measurement. The patient’s scores on the MDS-UPDRS III were 3 and 4 in the DBS OFF and ON setting, respectively. No medications were taken at the time of measurement.

For comparison, we present data from a female tremor-dominant idiopathic PD patient in her sixties in the Med ON state (first disease manifestation approximately 8 years ago at the time of participation; DBS system implanted 3 years ago; UPDRS Part III: Med OFF/DBS OFF: 51, Med ON/DBS ON: 18, Med OFF/DBS ON: 42). Both patients were implanted with a Medtronic Percept PC (Medtronic Inc., Minneapolis, MN, USA), capable of measuring local field potentials (LFPs) from the implanted DBS leads. DBS surgery was performed at the department of Functional Neurosurgery and Stereotaxy of the University Hospital Düsseldorf in adherence to standard procedures.

Both patients gave their written informed consent to participate in the study, according to the declaration of Helsinki. The study was approved by the Ethics Committee of the Medical Faculty of Heinrich Heine University Düsseldorf. Both patients consented to publication.

### Recordings and stimulation

MEG was measured using a 306-channel MEG system (VectorView, MEGIN, Espoo, Finland) with a sampling rate of 2 kHz. We additionally monitored horizontal and vertical ocular activity using electrooculography (EOG). Muscular activity was recorded via electromyography (EMG), with EMG surface electrodes placed on the patients’ right and left forearms, referenced to EMG electrodes on the wrist. Additional surface electrodes were placed on the left chest to track the electrocardiogram (ECG), above the implanted stimulator, as well as on the neck above the subcutaneous extension to record the DBS artifact, with reference electrodes positioned over the cervical vertebrae. We performed a 5 min resting-state recording in DBS OFF and subsequently applied monopolar, unilateral DBS using the second ring from the bottom (ring 1) at 130 Hz for 5 min in each hemisphere (amplitude: 1.2 mA; pulse width: 60 μs). We recorded bipolar LFPs with the Percept system in the BrainSense streaming mode from the rings above and below (0 and 2). Both patients additionally participated in a Go/NoGo task (see below). No stimulation was applied during the task.

### Electrode localization

DBS electrode localizations (Fig. 1) were performed with the advanced processing pipeline in Lead-DBS v3.1 (lead-dbs.org) ^25^. Briefly, postoperative CT images were linearly co-registered with pre-operative MRIs (T1 and T2) using advanced normalization tools ANTs; stnava.github.io/ANTs/; ^26^. If necessary, co-registrations were reviewed and refined. Brain shift corrections were performed using Lead-DBS standard tools. We used all preoperative volumes to estimate a precise multispectral normalization to ICBM 2009b NLIN asymmetric (“MNI”) space ^27^ using the ANTs SyN Diffeomorphic Mapping ^28^ with the preset “effective: low variance default + subcortical refinement.” The reconstruction of DBS contacts was performed manually or using the PaCER method ^29^. Atlas segmentations are based on the DISTAL atlas ^30^. Finally, using the Lead group toolbox, visualizations of the electrode reconstructions were generated for both patients ^31^.

**Figure 1.**
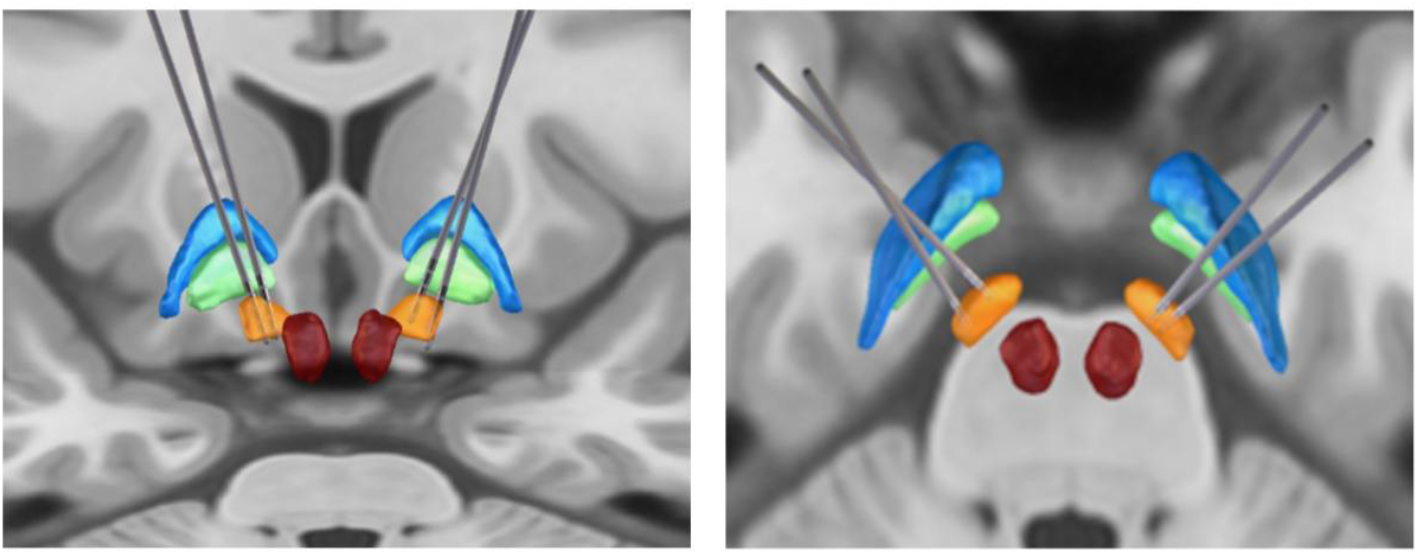
Electrode localization. Left: front view. Right: top view. Subthalamic nucleus: orange, external pallidum: blue, internal pallidum: green, red nucleus: red. The electrodes of the OCD patient are the ones that are positioned more medial on the level of the STN.

### Paradigm

Both patients completed a visually cued Go/NoGo task (OCD patient: 4 blocks; PD patient: 3 blocks; 120 trials per block) while seated in the MEG scanner (Fig. 2). Visual stimuli were presented using the software PsychoPy (version 2023.2.3) in Python (3.12.0). Individual reaction time was estimated at the beginning of the experiment through a sequence of Go trials. In the main experiment, each trial began with a black fixation cross, lasting 500 ms, and ended with feedback (on screen for 1 s). Following the fixation cross, we presented the outlines of a bar in either horizontal or vertical orientation (cue). After 500 ms, the bar acquired either an orange or a blue color fill, corresponding to the Go stimulus or the NoGo stimulus, respectively. In case of Go, the patient had to press a button with the right index finger as fast as possible (time limit: individual reaction time + 2 SD). In case of NoGo, the patient was instructed to withhold any response. The NoGo stimulus was on screen for the individual reaction time + 4 SD. The orientation of the bar predicted the upcoming stimulus (Go or NoGo), i.e. each orientation was preferentially paired with a particular color, and this preference needed to be learned on task. We refer to the more common pairing as congruent trials, and to the less common pairing as incongruent trials. The distribution of trials was: 55% congruent Go, 12.5% incongruent Go, 20% congruent NoGo and 12.5% incongruent NoGo.

**Figure 2.**
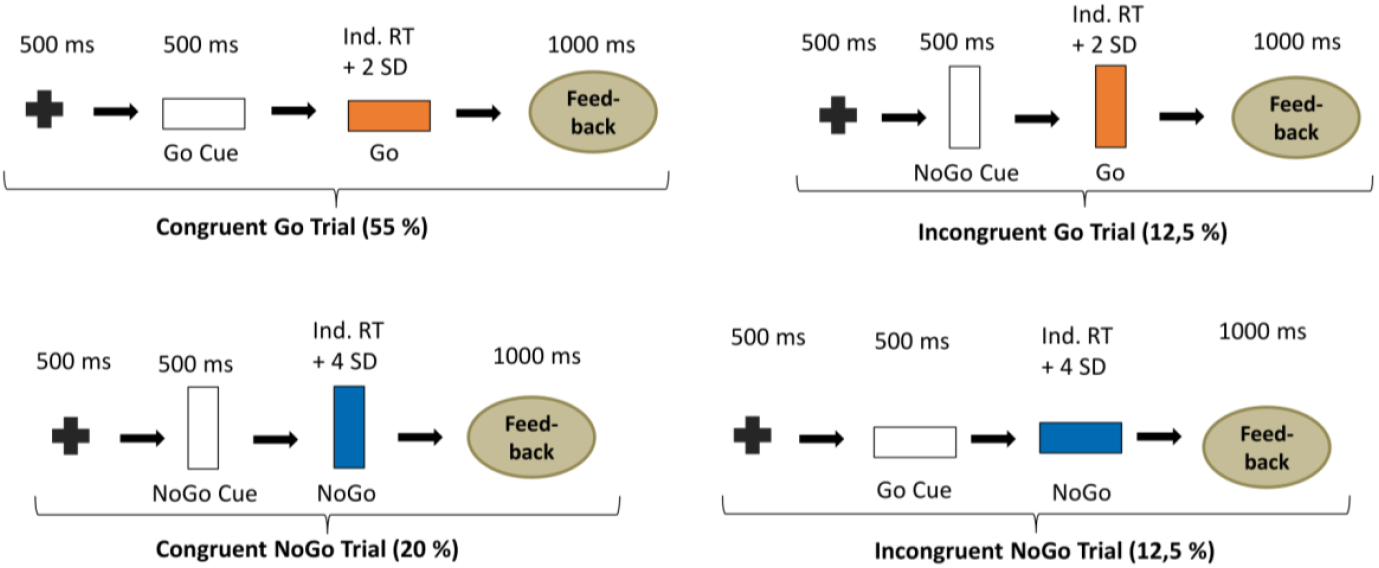
Cued Go/NoGo task. The association between bar orientation and expected instruction (here: horizontal – likely Go, vertical – likely NoGo) and between colour fill and instruction (here: orange – Go, blue – NoGo) was counterbalanced across subjects. Trial frequencies are noted below each trial type.

### Data analysis

Data were analyzed using MATLAB R2019b (The Mathworks, Natick, Massachusetts, USA) and the toolbox FieldTrip ^32^, as well as Python (Version 3.12) and the fitting oscillations and one over F (FOOOF) toolbox ^33^.

#### Preprocessing

We first visually identified noisy channels and subsequently applied temporal Signal Space Separation to the MEG data ^34^. The MEG data was downsampled to 250 Hz to match the sampling rate of the LFP data. We then applied a high-pass finite impulse response filter with a cut-off frequency of 1 Hz to remove low-frequency drifts and a low pass filter with a cut-off frequency of 100 Hz to both the MEG and LFP data to ease the detection of cardiac artifacts (see below).

We switched DBS on briefly at the beginning and at the end of each measurement, resulting in DBS artifacts, which we used for temporal alignment of MEG and LFP signals ^35^. Given the presence of strong cardiac artifacts in the resting-state STN LFP data when DBS was ON, this initial alignment could be improved further in a second step based on the ECG. First, the ECG signal was *z*-scored over the entire recording. Then, the R-peaks, features of the prominent QRS waveform in ECG signals, were identified using the Matlab function *findpeaks()* with two criteria: the peak height exceeded the mean signal level by 2.5 standard deviations, and the interval between successive peaks was at least 500 ms. Next, we defined epochs centered on the R-peak and averaged the epochs to obtain a mean QRS waveform for both LFP and ECG. Finally, we computed the cross-covariance between the two versions of the heartbeat and finetuned the initial alignment by correcting any delay visible in the cross-correlogram.

#### Spectral analysis and time-frequency analysis

LFP power and LFP-MEG coherence were computed using Welch’s method in combination with a Hanning taper ^32^. For LFP power, we isolated the oscillatory components from the aperiodic background using the FOOOF toolbox ^33^.

For the Go/NoGo task data, we source-reconstructed the activity of left and right primary motor cortices (M1, hand-knob) using Linearly Constrained Minimum Variance beamforming ^36^. Time-frequency spectra (2-45 Hz) were computed for STN and M1 bilaterally using a Hanning taper. As baseline, we used power averaged over all trial types (i.e. congruent/incongruent Go/NoGo trials) and all time points within those trials. In Fig. 4, we pooled congruent and incongruent trials, as we did not observe any effect of congruency.

#### Source reconstruction

To localize the sources of STN-cortex beta coherence, we first co-registered the pre-operative T1-weighted MRI scan with the MEG coordinate system. Using the segmented MRI, a forward model was generated based on a single-shell realistic head model ^37^. Beamformer grid points covered the whole brain, with their coordinates standardized to Montreal Neurological Institute (MNI) space. *Dynamic Imaging of Coherent Sources* (DICS) ^38^ was applied to beta-band LFP-MEG cross-spectral densities (13-30 Hz). To contrast the DBS ON and OFF conditions (Fig. 3), we averaged the source images for left and right stimulation and subtracted the DBS off image from the average.

**Figure 3.**
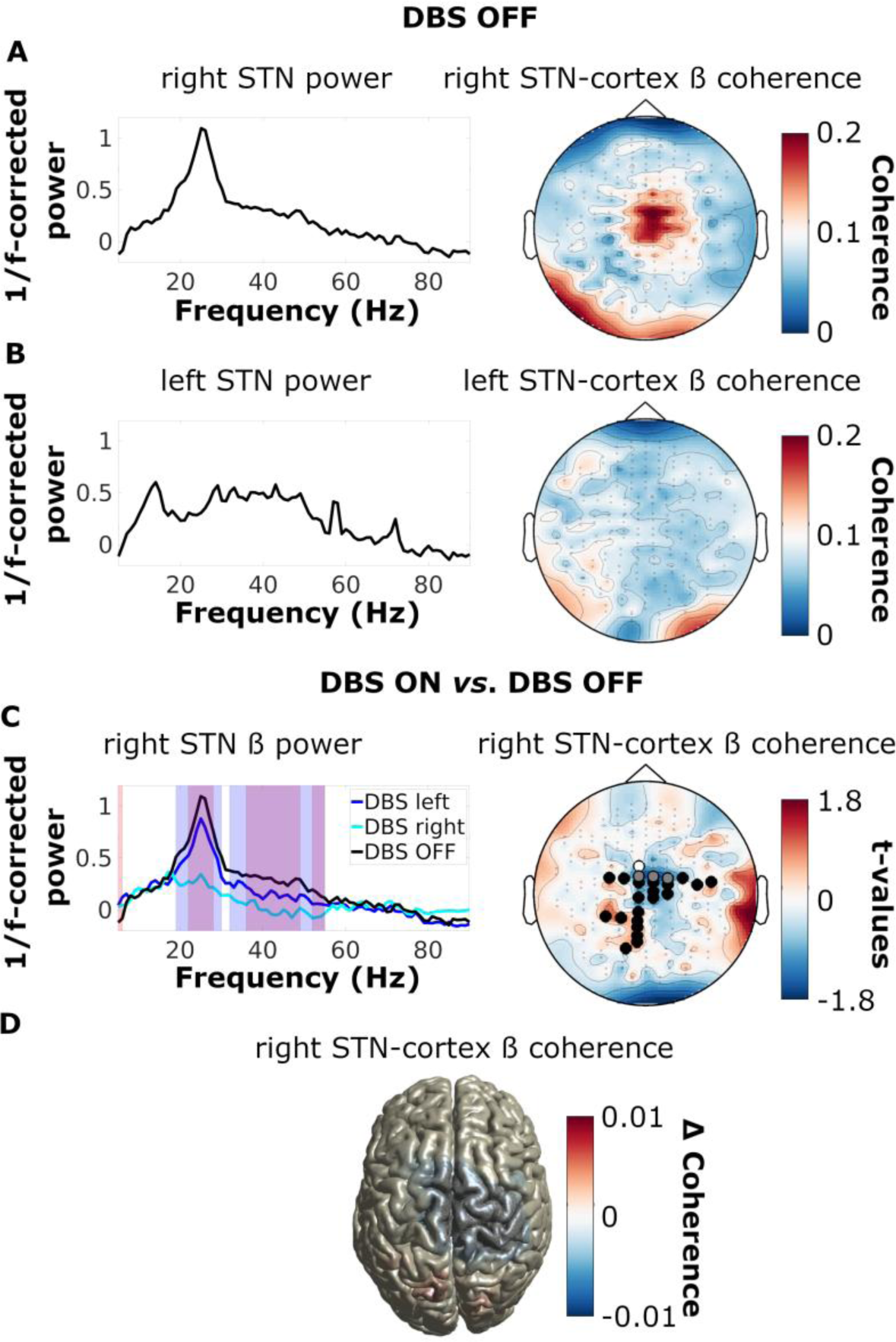
DBS reduced beta power and beta coherence in the OCD patient. Log10-transformed resting-state power spectra (aperiodic fit subtracted) and topographies of beta (13-30 Hz) STN-MEG sensor coherence for the right **(A)** and left **(B)** STN. **(C; left)** Log10-transformed resting-state power spectra (aperiodic fit subtracted) of the right STN during DBS OFF and during right and left DBS ON. Significant differences for right DBS ON *vs*. OFF: blue shade; left DBS ON *vs*. OFF: red shade; overlapping clusters: purple shade. **(C; right)** Topography of coherence between the right STN and the MEG sensors during right DBS ON *vs*. OFF and left DBS ON *vs*. OFF. Channels significantly modulated by right DBS are marked in black (left DBS: white, overlap: grey). **(D)** Source-localized contrast between DBS ON and DBS OFF (left and right DBS averaged).

#### Statistical analysis

Differences in power and coherence between DBS ON and OFF in resting-state, and power differences between Go and NoGo trials were identified through cluster-based permutation tests ^39^. We performed 1000 permutations, and used a cluster defining threshold of 0.05 (two-sided test) and an alpha level of 0.025. The cluster statistic was defined as the sum of *t*-values within a cluster.

### Data availability

Data can be made available in anonymized form upon reasonable request.

## Results

### DBS reduced STN beta power and STN-cortex beta coherence in OCD

When analyzing the resting-state data of the OCD patient, we observed a prominent peak in the beta-band for right STN power and for coherence between right STN and right sensorimotor cortex (Fig. 3A). The beta power peak was reduced by stimulation of the left (*t*_clustersum_ ≤ −10.775, *p* < 0.001) and particularly of the right (*t*_clustersum_ ≤ −95.168, *p* < 0.001) STN (Fig. 3C). Similarly, the beta peak in STN-sensorimotor cortex coherence was suppressed by DBS of the right (*t*_clustersum_ ≤ −14.258, *p* ≤ 0.02) or left STN (*t*_clustersum_ ≤ −14.704, *p* ≤ 0.002; Fig. 3C, D). Detailed statistical results can be found in Supplemental Tables 1-2.

### Task-related modulations of subthalamic oscillations differed between the OCD and the PD patient

Surprised by how closely the resting-state patterns of the OCD patient resembled those reported for PD, we wondered whether we would find a more distinct oscillatory signature in a task. Thus, we had the OCD patient perform a Go/NoGo task and compared the recordings to a PD patient measured with the same setup.

On the cortical level, the responses were rather similar (Fig. 4). In Go trials, we observed movement-related beta suppression after the Go stimulus, followed by a beta rebound. In NoGo trials, the suppression was interrupted by an early increase in beta power, differentiating response inhibition from execution (OCD M1: *t*_clustersum_ = −561.202, *p* < 0.001; PD M1: *t*_clustersum_ = −711.0559, *p* < 0.001). On the level of the STN, the beta-dominated pattern seen in cortex repeated in the PD patient (*t*_clustersum_ = −312.723, *p* < 0.001), but appeared to be shifted in frequency in the OCD patient, with differences arising in the theta band in left (Fig. 4) and right STN (Supplemental Fig. 1). Specifically, NoGo trials were associated with higher theta power than Go trials (*t*_clustersum_ = −333.027, *p* < 0.001; Fig. 4A). Details are reported in Supplemental Tables 3-4.

**Figure 4.**
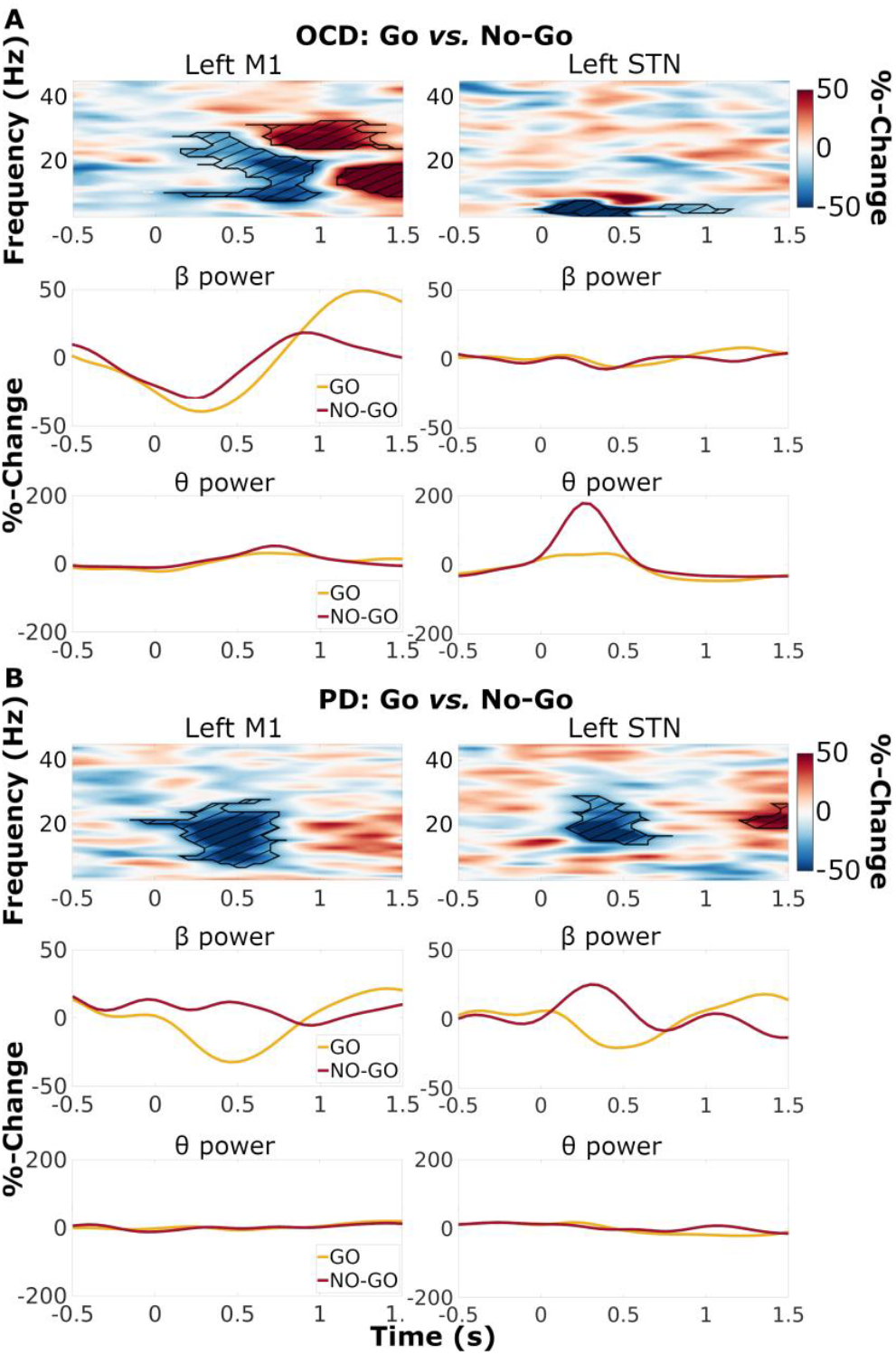
Modulations of left subthalamic power associated with response inhibition differed between the OCD and the PD patient. Top: time-frequency power spectra, contrast between Go and NoGo trials (pooled over cue types), for the OCD patient **(A)** and the PD patient **(B)**. The difference between baseline-corrected Go trials and baseline-corrected NoGo trials is color-coded. Significant differences are marked by hatched lines within contours. Bottom: band-average power time course, for beta (13-30 Hz), and theta (3-8 Hz) frequencies.

## Discussion

DBS in OCD is rare and its effects on neural oscillations and their synchronization across basal ganglia cortex loops are underexplored. Here, we demonstrate the existence of prominent beta oscillations, synchronized across STN and motor cortex in the resting-state, in a single OCD patient. Interestingly, DBS suppressed these oscillations, as reported previously for PD patients ^7,9,40^. Notably, we did not observe any DBS-entrained gamma activity in the OCD patient at half the stimulation frequency, in line with the idea that it is related to dopaminergic medication intake ^41^.

By providing a non-movement disorder control, this paper makes an important contribution to the discussion on the link between two major effects of DBS: the dampening of beta oscillations and the concurrent improvement of motor symptoms. Our results clearly indicate that these two effects can (but need not) dissociate, with beta-suppression occurring in the absence of any changes in motor performance. We thus conclude that DBS-responsive beta oscillations are not necessarily a sign of PD. The fact that they do occur in several diseases suggests that they might relate to physiological functions ^42^, such as somatosensory processing, the integration of sensory feedback with existing knowledge ^43^, and the maintenance of current motor/cognitive output ^44^. Alternatively, they might represent a common feature of PD and OCD, such as a high level of inhibition, arising either as a consequence of neurodegeneration (PD) or from volitional processes such as withstanding compulsions (OCD). The STN is likely involved in either process, as it integrates cognitive, limbic and motor processes ^42^, with beta oscillations occurring in both motor-^17^ and non-motor ^11,45^ regions.

In contrast to the resting-state recordings, the Go/NoGo task revealed an oscillatory pattern not familiar from the PD literature, which emphasizes the involvement of beta oscillations ^46^. In the OCD patient, however, we found the strongest responses in the theta band. This finding aligns with previous research linking altered theta activity to OCD symptoms ^16,17^, such as the inability to inhibit compulsive behaviors or deal with conflict, an established correlate of theta activity ^47^. To ensure that the deviation was not due to the methodology applied here, we repeated the experiment in a single PD patient. As expected, we observed marked modulations of STN beta power, possibly reflecting the disease’s characteristic overactivity of STN-cortical pathways. Notably, motor cortex exhibited task-related beta modulations in both patients. This might have led to corresponding beta-band modulations of subthalamic activity in PD only, due to insufficient shielding of the STN from motor cortical drive ^3^.

Of course, these ideas need to be tested in group studies. Case studies are limited by design, particularly when comparing DBS patients with different electrode placements (Fig. 1). We cannot rule out that a more anteromedial vs. dorsolateral electrode placement explains the spectral shift observed here (beta modulation in PD, theta modulation in OCD). Yet, both of the subthalamic compartments sampled here feature both theta and beta oscillations ^45,48^, suggesting that the spectral shift is due to the disease rather than the subthalamic compartment.

In summary, this case study illustrates how a task may uncover disease-specific STN oscillations that are not apparent in resting-state. This aligns with functional MRI research, demonstrating that task-based connectivity patterns contain more behaviorally relevant information than resting-state connectivity ^49,50^. Importantly, our study proves that DBS-responsive beta oscillations exist in non-movement disorders, demonstrating that these oscillations are not necessarily a sign of motor impairment.

## References

1. Brown P, Oliviero A, Mazzone P, Insola A, Tonali P, Di Lazzaro V. Dopamine dependency of oscillations between subthalamic nucleus and pallidum in Parkinson’s disease. J Neurosci. 2001;21(3):1033–1038.

2. Hirschmann J, Özkurt TE, Butz M, et al. Distinct oscillatory STN-cortical loops revealed by simultaneous MEG and local field potential recordings in patients with Parkinson’s disease. NeuroImage. 2011;55(3):1159–1168.

3. Oswal A, Cao C, Yeh C-H, et al. Neural signatures of hyperdirect pathway activity in Parkinson’s disease. Nature Communications. 2021;12(1):5185.

4. Kühn AA, Tsui A, Aziz T, et al. Pathological synchronisation in the subthalamic nucleus of patients with Parkinson’s disease relates to both bradykinesia and rigidity. Experimental Neurology. 2009;215(2):380–387.

5. Lofredi R, Tan H, Neumann W-J, et al. Beta bursts during continuous movements accompany the velocity decrement in Parkinson’s disease patients. Neurobiol Dis. 2019;127:462–471.

6. Muthuraman M, Bange M, Koirala N, et al. Cross-frequency coupling between gamma oscillations and deep brain stimulation frequency in Parkinson’s disease. Brain. 2020;143(11):3393–3407.

7. Mathiopoulou V, Lofredi R, Feldmann LK, et al. Modulation of subthalamic beta oscillations by movement, dopamine, and deep brain stimulation in Parkinson’s disease. npj Parkinson’s Disease. 2024;10(1):77.

8. Wilkins KB, Petrucci MN, Lambert EF, et al. Beta burst-driven adaptive deep brain stimulation improves gait impairment and freezing of gait in Parkinson’s disease. medRxiv : the preprint server for health sciences. 2024.

9. Abbasi O, Hirschmann J, Storzer L, et al. Unilateral deep brain stimulation suppresses alpha and beta oscillations in sensorimotor cortices. NeuroImage. 2018;174:201–207.

10. American Psychiatric Association. Diagnostic and statistical manual of mental disorders 5ed: American Psychiatric Publishing; 2013.

11. Wojtecki L, Hirschmann J, Elben S, et al. Oscillatory coupling of the subthalamic nucleus in obsessive compulsive disorder. Brain. 2017;140(9):e56.

12. Qing X, Gu L, Li D. Abnormalities of localized connectivity in obsessive-compulsive disorder: A voxel-wise meta-analysis. Frontiers in Human Neuroscience. 2021;15:739175.

13. Beucke JC, Sepulcre J, Talukdar T, et al. Abnormally high degree connectivity of the orbitofrontal cortex in obsessive-compulsive disorder. JAMA Psychiatry. 2013;70(6):619–629.

14. Smith EE, Schüller T, Huys D, et al. A brief demonstration of frontostriatal connectivity in OCD patients with intracranial electrodes. NeuroImage. 2020;220:117138.

15. Horn A, Li N, Meyer GM, Gadot R, Provenza NR, Sheth SA. Deep Brain Stimulation response circuits in Obsessive Compulsive Disorder. Biological Psychiatry.

16. Bastin J, Polosan M, Piallat B, et al. Changes of oscillatory activity in the subthalamic nucleus during obsessive-compulsive disorder symptoms: Two case reports. Cortex. 2014;60:145–150.

17. Rappel P, Marmor O, Bick AS, et al. Subthalamic theta activity: a novel human subcortical biomarker for obsessive compulsive disorder. Transl Psychiatry. 2018;8(1):118.

18. Desarkar P, Sinha VK, Jagadheesan K, Nizamie SH. Subcortical functioning in obsessive-compulsive disorder: an exploratory EEG coherence study. The World Journal of Biological Psychiatry. 2007;8(3):196–200.

19. Lee I, Kim KM, Lim MH. Theta and gamma activity differences in obsessive-compulsive disorder and panic disorder: Insights from resting-state eeg with eLORETA. Brain sciences. 2023;13(10).

20. Perera MPN, Mallawaarachchi S, Bailey NW, Murphy OW, Fitzgerald PB. Obsessive-compulsive disorder (OCD) is associated with increased electroencephalographic (EEG) delta and theta oscillatory power but reduced delta connectivity. Journal of Psychiatric Research. 2023;163:310–317.

21. Figee M, Luigjes J, Smolders R, et al. Deep brain stimulation restores frontostriatal network activity in obsessive-compulsive disorder. Nature Neuroscience. 2013;16(4):386–387.

22. Schwabe K, Alam M, Saryyeva A, et al. Oscillatory activity in the BNST/ALIC and the frontal cortex in OCD: Acute effects of DBS. Journal of Neural Transmission. 2021;128(2):215–224.

23. Koh MJ, Seol J, Kang JI, et al. Altered resting-state functional connectivity in patients with obsessive–compulsive disorder: A magnetoencephalography study. International Journal of Psychophysiology. 2018;123:80–87.

24. Xiong B, Wen R, Gao Y, Wang W. Longitudinal changes of local field potential oscillations in nucleus accumbens and anterior limb of the internal capsule in obsessive-compulsive disorder. Biological Psychiatry. 2023;93(11):e39–e41.

25. Horn A, Kühn AA. Lead-DBS: A toolbox for deep brain stimulation electrode localizations and visualizations. NeuroImage. 2015;107:127–135.

26. Avants BB, Tustison NJ, Song G, Cook PA, Klein A, Gee JC. A reproducible evaluation of ANTs similarity metric performance in brain image registration. NeuroImage. 2011;54(3):2033–2044.

27. Fonov V, Evans AC, Botteron K, Almli CR, McKinstry RC, Collins DL. Unbiased average age-appropriate atlases for pediatric studies. NeuroImage. 2011;54(1):313–327.

28. Avants BB, Epstein CL, Grossman M, Gee JC. Symmetric diffeomorphic image registration with cross-correlation: Evaluating automated labeling of elderly and neurodegenerative brain. Medical Image Analysis. 2008;12(1):26–41.

29. Husch A, V. Petersen M, Gemmar P, Goncalves J, Hertel F. PaCER - A fully automated method for electrode trajectory and contact reconstruction in deep brain stimulation. NeuroImage: Clinical. 2018;17:80–89.

30. Ewert S, Plettig P, Li N, et al. Toward defining deep brain stimulation targets in MNI space: A subcortical atlas based on multimodal MRI, histology and structural connectivity. NeuroImage. 2018;170:271–282.

31. Treu S, Strange B, Oxenford S, et al. Deep brain stimulation: Imaging on a group level. NeuroImage. 2020;219:117018.

32. Oostenveld R, Fries P, Maris E, Schoffelen JM. FieldTrip: Open source software for advanced analysis of MEG, EEG, and invasive electrophysiological data. Computational Intelligence and Neuroscience. 2011;2011(1):156869.

33. Donoghue T, Haller M, Peterson EJ, et al. Parameterizing neural power spectra into periodic and aperiodic components. Nature Neuroscience. 2020;23(12):1655–1665.

34. Taulu S, Simola J. Spatiotemporal signal space separation method for rejecting nearby interference in MEG measurements. Phys Med Biol. 2006;51(7):1759–1768.

35. Hnazaee MF, Sure M, O’Neill GC, et al. Combining magnetoencephalography with telemetric streaming of intracranial recordings and deep brain stimulation—A feasibility study. Imaging Neuroscience. 2023;1:1–22.

36. Van Veen BD, Buckley KM. Beamforming: A versatile approach to spatial filtering. IEEE ASSP Magazine. 1988;5(2):4–24.

37. Nolte G. The magnetic lead field theorem in the quasi-static approximation and its use for magnetoencephalography forward calculation in realistic volume conductors. Phys Med Biol. 2003;48(22):3637–3652.

38. Gross J, Kujala J, Hämäläinen M, Timmermann L, Schnitzler A, Salmelin R. Dynamic imaging of coherent sources: Studying neural interactions in the human brain. Proceedings of the National Academy of Sciences. 2001;98(2):694–699.

39. Maris E, Oostenveld R. Nonparametric statistical testing of EEG- and MEG-data. Journal of neuroscience methods. 2007;164(1):177–190.

40. Oswal A, Beudel M, Zrinzo L, et al. Deep brain stimulation modulates synchrony within spatially and spectrally distinct resting state networks in Parkinson’s disease. Brain. 2016;139(5):1482–1496.

41. Colombo A, Bernasconi E, Alva L, et al. Finely tuned γ tracks medication cycles in Parkinson’s disease: An ambulatory brain-sense study. Movement Disorders. 2025.

42. Accolla EA, Horn A, Herrojo-Ruiz M, Neumann W-J, Kühn AA. Reply: Oscillatory coupling of the subthalamic nucleus in obsessive compulsive disorder. Brain. 2017;140(9):e57–e57.

43. Barone J, Rossiter HE. Understanding the role of sensorimotor beta oscillations. Front Syst Neurosci. 2021;15(51).

44. Engel A, Fries P. Beta-band oscillation - signalling the status quo? Current Opinion in Neurobiology. 2010;20(2):156–165.

45. Accolla EA, Herrojo Ruiz M, Horn A, et al. Brain networks modulated by subthalamic nucleus deep brain stimulation. Brain. 2016;139(Pt 9):2503–2515.

46. Kühn AA, Williams D, Kupsch A, et al. Event-related beta desynchronization in human subthalamic nucleus correlates with motor performance. Brain. 2004;127(4):735–746.

47. Zavala B, Brittain J-S, Jenkinson N, et al. Subthalamic nucleus local field potential activity during the Eriksen Flanker Task reveals a novel role for theta phase during conflict monitoring. J Neurosci. 2013;33(37):14758.

48. Olson JW, Nakhmani A, Irwin ZT, et al. Cortical and subthalamic nucleus spectral changes during limb movements in Parkinson’s disease patients with and without dystonia. Movement Disorders. 2022;37(8):1683–1692.

49. Zhao W, Makowski C, Hagler DJ, et al. Task fMRI paradigms may capture more behaviorally relevant information than resting-state functional connectivity. NeuroImage. 2023;270:119946.

50. Elliott ML, Knodt AR, Cooke M, et al. General functional connectivity: Shared features of resting-state and task fMRI drive reliable and heritable individual differences in functional brain networks. NeuroImage. 2019;189:516–532.

