## Supplemental Materials for "Deep brain stimulation-responsive subthalamo-cortical coupling in obsessive-compulsive disorder"

### Supplemental Results

##### Supplemental Table 1: Effects of left hemispheric DBS on STN power.

| Freq. start [Hz] | Freq. end [Hz] | cluster statistic | *p*-value |
|---|---|---|---|
| 36 | 49 | -39.547 | <0.001 |
| 22 | 28 | -22.863 | <0.001 |
| 52 | 55 | -10.775 | <0.001 |
| 5 | 6 | 7.3950 | 0.010 |

The results from the cluster-based permutation test describe the effects of left hemispheric DBS on right STN power (spectral test). The cluster statistic refers to the sum of t-values within a cluster. The sign of the statistic indicates whether STN power was increased or reduced by DBS. Only clusters significant in a two-sided test are listed.

##### Supplemental Table 2: Effects of right hemispheric DBS on STN power.

| Freq. start [Hz] | Freq. end [Hz] | cluster statistic | *p*-value |
|---|---|---|---|
| 32 | 55 | -130.335 | <0.001 |
| 19 | 30 | -95.168 | <0.001 |

The results from the cluster-based permutation test describe the effects of right hemispheric DBS on right STN power (spectral test). The cluster statistic refers to the sum of t-values within a cluster. The sign of the statistic indicates whether STN power was increased or reduced by DBS. Only clusters significant in a two-sided test are listed.

**Supplemental Table 3: Effects of left hemispheric DBS on STN-cortex coherence.**

| location | cluster statistic | *p*-value |
|---|---|---|
| Bilateral sensorimotor cortices | -14.704 | 0.002 |

The results from the cluster-based permutation test describe the effects of left hemispheric DBS on beta coherence between right STN and cortex (spatial test, sensor level). The cluster statistic refers to the sum of t-values within a cluster. The sign of the statistic indicates whether coherence was increased or reduced by DBS. Only clusters significant in a two-sided test are listed.

**Supplemental Table 4: Effects of right-hemispheric DBS on STN-cortex coherence.**

| location | cluster statistic | *p*-value |
|---|---|---|
| Left Sensorimotor cortex | -104.829 | <0.001 |
| Left parietal cortex | 60.482 | <0.001 |
| Right Sensorimotor cortex | -24.462 | 0.004 |

The results from the cluster-based permutation test describe the effects of right hemispheric DBS on beta coherence between right STN and cortex (spatial test, sensor level). The cluster statistic refers to the sum of t-values within a cluster. The sign of the statistic indicates whether coherence was increased or reduced by DBS. Only clusters significant in a two-sided test are listed.

**Supplemental Table 5: Left M1 power during Go *vs.* NoGo trials in OCD.**

| frequencies [Hz] | time points [s] | cluster statistic | *p*-value |
|---|---|---|---|
| 7.568 - 28.809 | 0.052 – 1.000 | -561.202 | <0.001 |
| 23.682 - 32.471 | 0.548 - 1.500 | 316.069 | <0.001 |
| 8.789 - 18.799 | 1.100 - 1.500 | 221.342 | <0.001 |

The results from cluster-based permutation test describe the effects of trial type (Go or NoGo) on time-resolved power in the left M1 of the OCD patient (time-frequency test). The cluster statistic refers to the sum of t-values within a cluster. The sign of the statistic indicates whether baseline-corrected power increased or decreased in Go compared to NoGo trials. Only clusters significant in a two-sided test are listed.

**Supplemental Table 6: Left STN power during Go *vs.* NoGo trials in OCD.**

| frequencies [Hz] | time points [s] | cluster statistic | *p*-value |
|---|---|---|---|
| 2.441 - 7.568 | -0.048 - 1.152 | -333.027 | <0.001 |

The results from cluster-based permutation test describe the effects of trial type (Go or NoGo) on time-resolved power in the left STN of the OCD patient (time-frequency test). The cluster statistic refers to the sum of t-values within a cluster. The sign of the statistic indicates whether baseline-corrected power increased or decreased in Go compared to NoGo trials. Only clusters significant in a two-sided test are listed.

**Supplemental Table 7: Left M1 power during Go *vs.* NoGo trials in PD.**

| frequencies [Hz] | time points [s] | cluster statistic | *p*-value |
|---|---|---|---|
| 6.348 - 27.588 | -0.148 - 0.800 | -711.056 | <0.001 |

The results from cluster-based permutation test describe the effects of trial type (Go or NoGo) on time-resolved power in the left M1 of the PD patient (time-frequency test). The cluster statistic refers to the sum of t-values within a cluster. The sign of the statistic indicates whether baseline-corrected power increased or decreased in Go compared to NoGo trials. Only clusters significant in a two-sided test are listed.

**Supplemental Table 8: Left STN power during Go *vs.* NoGo trials in PD.**

| | | | |
|---|---|---|---|
| 13.672 - 28.809 | 0.152 - 0.800 | -312.723 | <0.001 |
| 18.799 - 26.367 | 1.200 - 1.500 | 81.775 | 0.004 |

The results from cluster-based permutation test describe the effects of trial type (Go or NoGo) on time-resolved power in the left STN of the PD patient (time-frequency test). The cluster statistic refers to the sum of t-values within a cluster. The sign of the statistic indicates whether baseline-corrected power increased or decreased in Go compared to NoGo trials. Only clusters significant in a two-sided test are listed.

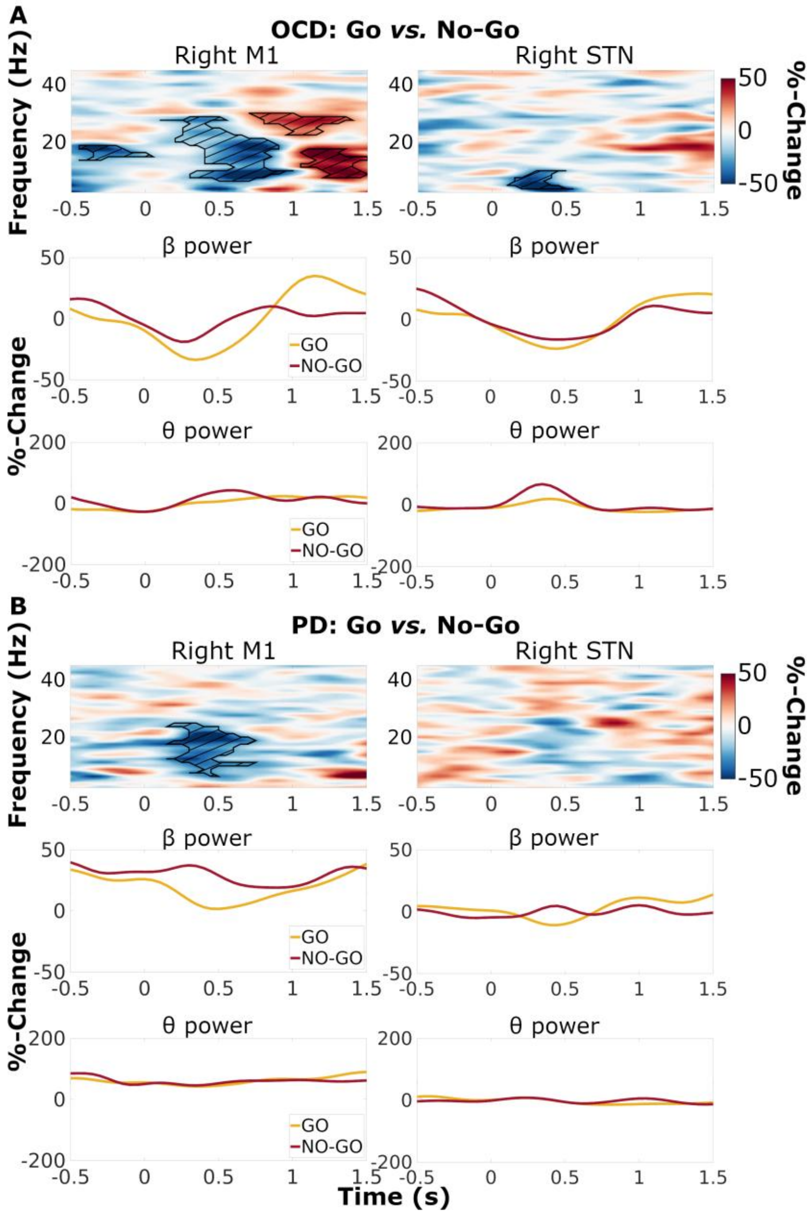


**Supplemental Figure 1: Modulations of right subthalamic power associated with response inhibition differed between the OCD and the PD patient**. Top: time-frequency power spectra, contrast between Go and NoGo trials (pooled over cue types), for the OCD patient **(A)** and the PD patient **(B)**. The difference between baseline-corrected Go trials (% change) and baseline-corrected NoGo trials (% change) is color-coded. Significant differences are marked by hatched lines within contours. Bottom: band-average power time course, for beta (13-30 Hz), and theta (3-8 Hz) frequencies.
